# Inflammatory, but not respiratory symptoms, associated with ongoing upper airway viral replication in outpatients with uncomplicated COVID-19

**DOI:** 10.1101/2021.03.05.21253011

**Authors:** Karen B. Jacobson, Natasha Purington, Julie Parsonnet, Jason Andrews, Vidhya Balasubramanian, Hector Bonilla, Karlie Edwards, Manisha Desai, Upinder Singh, Haley Hedlin, Prasanna Jagannathan

## Abstract

**Background:** The vast majority of SARS-CoV-2 infections are uncomplicated and do not require hospitalization, but contribute to ongoing transmission. Our understanding of the clinical course of uncomplicated COVID-19 remains limited.

**Methods:** We detailed the natural history of uncomplicated COVID-19 among 120 outpatients enrolled in a randomized clinical trial of Peginterferon Lambda. We characterized symptom trajectory and clusters using exploratory factor analysis, assessed predictors of symptom resolution and cessation of oropharyngeal viral shedding using Cox proportional hazard models, and evaluated associations between symptoms and viral shedding using mixed effects linear models.

**Results:** Headache, myalgias and chills peaked at day 4 after symptom onset; cough peaked on day 9. Two distinct symptom cluster trajectories were identified; one with mild, upper respiratory symptoms, and the other with more severe and prolonged inflammatory symptoms. The median time to symptom resolution from earliest symptom onset was 17 days (95% CI 14-18). Neither enrollment SARS-CoV-2 IgG levels (Hazard ratio [HR] 1.88, 95% CI 0.84-4.20) nor oropharyngeal viral load at enrollment (HR 1.01, 95% CI 0.98-1.05) were significantly associated with the time to symptom resolution. The median time to cessation of viral shedding was 10 days (95% CI 8-12), with higher SARS-CoV-2 IgG levels at enrollment associated with hastened resolution of viral shedding (HR 3.12, 95% CI 1.4-6.9, p=0.005). Myalgia, joint pains, and chills were associated with a significantly greater odds of oropharyngeal SARS-CoV-2 RNA detection.

**Conclusions:** In this outpatient cohort, inflammatory symptoms peaked early and were associated with ongoing SARS-CoV-2 replication. SARS-CoV-2 antibody levels were associated with more rapid viral shedding cessation, but not with time to symptom resolution. These findings have important implications for COVID-19 screening approaches and clinical trial design.

## Introduction

As of February 2021, 109 million individuals have been confirmed infected with SARS-CoV-2, and over 2.4 million have died.(1) Despite high mortality in the elderly and among those with medical comorbidities, the vast majority of individuals have uncomplicated disease, with only mild to moderate symptoms, and do not require hospitalization. However, these individuals contribute to ongoing transmission, and some develop prolonged symptoms and sustained disability.(2) A better understanding of the natural history of uncomplicated COVID-19 would help guide the development of therapeutics needed to shorten the duration of viral shedding, relieve symptoms, and prevent hospitalizations.

Most published reports of COVID-19 symptomatology are derived from hospitalized patients, in whom fever and shortness of breath are common (3-5). In contrast, outpatients with more mild disease report less fever, but hyposmia is more common(6, 7). Data on symptom trajectories and clusters is limited, as studies in outpatients have largely been limited to cross-sectional surveys. Furthermore, the relationship of these syndromes to viral shedding and SARS-CoV-2 antibody responses have not been described in detail.

In this study, we leveraged a cohort of participants with uncomplicated COVID-19 enrolled in a randomized, placebo-controlled trial of Peginterferon Lambda to perform a detailed examination of the symptomatology and natural history of uncomplicated SARS-CoV-2 infection. We aimed to 1) characterize symptom trajectories and clusters; 2) assess predictors of symptom resolution and cessation of oropharyngeal viral shedding; and 3) evaluate associations between symptoms and viral shedding.

## Methods

### Study setting and participants

SARS-CoV-2-infected outpatients were enrolled in a double-blind, placebo-controlled, phase 2 trial of Peginterferon Lambda-1a (Lambda, NCT04331899). Inclusion/exclusion criteria and the study protocol for the trial have been published.(8) Briefly, adults aged 18-75 years with an FDA emergency use authorized reverse transcription-polymerase chain reaction (RT-PCR) positive for SARS-CoV-2 within 72 hours prior to enrollment were eligible for study participation. Exclusion criteria included hospitalization, respiratory rate >20 breaths per minute, room air oxygen saturation <94%, pregnancy or breastfeeding, decompensated liver disease, recent use of investigational and/or immunomodulatory agents for treatment of COVID-19, and prespecified lab abnormalities.

### Study procedures

On the day of enrollment, participants completed a standardized history, including the duration of symptoms prior to enrollment, physical exam and bloodwork. Participants were randomized 1:1 to receive a single subcutaneous injection of Lambda or saline placebo and followed for 28 days. Participants completed a daily symptom questionnaire and recorded daily in-home measurements of temperature and oxygen saturation using study-provided devices (REDCap Cloud). In-person visits were conducted on Days 1, 3, 5, 7, 10, 14, 21, and 28 post-enrollment with assessment of symptoms and vitals, and collection of oropharyngeal swabs (FLOQ Swabs; Copan Diagnostics). Peripheral blood was collected for assessment of complete blood count, metabolic panel, and SARS-CoV-2 antibody levels against the SARS-CoV-2 spike receptor binding domain (RBD, Supplementary Methods).

### Statistical analysis

Analyses were conducted in R version 4.0.2(9) or Stata SE (version 14). Cohort demographics, baseline characteristics and distribution of symptom prevalence were summarized descriptively. Symptom duration prior to enrollment determined symptom start day. Symptoms reported pre-enrollment were assumed to have been continuously experienced from the first reported day until enrollment. Symptoms occurring more than 21 days prior to enrollment were deemed non-COVID related and excluded. Outcomes measured included symptom prevalence and duration, time to resolution of symptoms and cessation of viral shedding.

#### Symptom prevalence and duration

Differences in symptom prevalence and the proportion of participants reporting symptoms over time by treatment arm were tested using the Kruskal-Wallis rank sum test and Fisher’s exact test, respectively. Since duration of symptoms and duration of viral shedding were similar between treatment arms,(8) these tests confirmed adjustment for treatment arm was not needed in our symptom-specific adjusted models. We therefore included participants from both the Lambda and placebo arms in this analysis. Repeated measures of at-home temperature and oxygen saturation were modeled using linear regression models with robust standard errors and generalized estimating equations to account for repeated measures within participants.

#### Symptom clusters

Spearman’s correlation coefficient and exploratory factor analysis (EFA) were used to assess pairwise correlations between symptom severity and whether symptoms clustered at various days from symptom onset, respectively. EFA was carried out on symptoms reported at symptom onset and on days 4, 7, 10, 14, and 21 after symptom onset. Only symptoms with more than 5% overall study prevalence were chosen for examination (Supplementary Methods). Changes in factors derived and symptoms contributing to each factor were compared descriptively across time points of interest. The joint trajectory of symptoms was assessed to determine whether participant clusters persist over time (Supplementary Methods). Fisher’s exact test and the Kruskal-Wallis rank sum test were used to test for differences in baseline characteristics and demographics by cluster assignment.

#### Time to event analyses

Time to first symptom resolution among those reporting symptoms was defined in two ways: 1) as time from symptom onset, and 2) as time from enrollment, until the first day when no symptoms were reported, given that vital signs and laboratories were not measured prior to enrollment. Participants who did not experience resolution were censored on the day of their last reported symptom. Cox proportional hazards models were fit as a function of covariates of interest. Both models included age at randomization, sex, body mass index (BMI), and race/ethnicity. The time from enrollment model additionally included pre-enrollment symptom duration, oxygen saturation, heart rate, alanine transaminase (ALT), white blood count, lymphocyte count, oropharyngeal PCR result, and antibody levels at enrollment. The proportional hazards assumption was assessed by evaluating scaled Schoenfeld residuals.(10)

Time to cessation of viral shedding was defined as time from first recorded positive test (pre-enrollment) until first of two consecutive negative oropharyngeal tests. Participants who did not experience shedding cessation were censored at the time of their last positive test or Day 1 if no positive test was observed. Cox proportional hazards model were fit as a function of all demographic and vital/lab measurements at enrollment described above, excluding pre-enrollment symptom duration.

For all models, crude and adjusted hazard ratios, 95% confidence intervals, and Kaplan-Meier estimates were reported.

#### Associations between viral detection and symptomology

Mixed effect logistic regression models were fit to PCR result (positive vs negative) as a function of symptom reported (yes vs no), days from randomization, continuous age at randomization, sex, treatment arm, and a random effect for participant to account for repeated measures over time within a participant. This model was fit to each of the 16 symptoms. Days from enrollment was used in order to align the longitudinal PCR and symptom data. Only days when both the symptom questionnaire and PCR results were available were included. Odds ratios, 95% CIs, and p-values for symptom were reported from each model.

#### Additional analyses

Fisher’s exact test and the Kruskal-Wallis rank sum test were used to test for differences in baseline characteristics and demographics by development of antibodies at Day 28 among those negative at baseline.

P-values <0.05 were considered statistically significant. As these were secondary analyses, we did not adjust for multiplicity, but instead consider our findings to be hypothesis generating.

## Results

### Study participants

We enrolled 120 participants between April 25 and July 17, 2020 (Figure S1). The median age was 36 years, 50 participants (41.7%) were female, and 75 (62.5%) were LatinX ethnicity (Table 1). At enrollment, 3 (2.5%) of participants had a temperature >100.4° F, and 49 (40.8%) were SARS-CoV-2 IgG seropositive. Antibody level at enrollment was positively associated with day since symptom onset (Figure S2). Overall, 110 participants (91.7%) completed 28 days of follow up, and the proportion of missing follow-up visits was 44/960 (4.6%).

**Table 1.** Baseline demographics and clinical characteristics

| **Characteristic** | **Overall**<br>**(N=120)** |
| --- | --- |
| **Age in years**, median (range) | 36 (18-71) |
| **Female**, n (%) | 70 (58.3%) |
| **Race / Ethnicity**, n (%) |  |
| Latinx | 75 (62.5%) |
| White | 33 (27.5%) |
| Asian | 7 (5.8%) |
| Native Hawaiian or other Pacific Islander | 2 (1.7%) |
| Unknown | 2 (1.7%) |
| More than one race | 1 (0.8%) |
| **BMI at enrollment (kg/m<sup>2</sup>)**, median (IQR) | 27.7 (24.9-32.0) |
| **Vital signs at enrollment** |  |
| Temperature >100.4F, n (%) | 3 (2.5%) |
| Oxygen saturation, median (IQR) | 98 (3) |
| **Laboratory values at enrollment**, median (IQR) |  |
| White blood cell (WBC) count, cells/μl | 5.5 (4.1-7.1) |
| Absolute lymphocyte count (ALC), cells/μl | 1.5 (1.2-2.2) |
| Aspartate aminotransferase, IU/L | 30 (25-41) |
| Alanine aminotransferase, IU/L | 31.5 (22-50.3) |
| **Seropositivity (IgG) at enrollment**, n (%) | 49 (40.8%) |

BMI=Body Mass Index. IQR=interquartile range.

### Symptom prevalence and trajectories

The most prevalent symptoms over the course of 28 days of follow up were fatigue (n=95, 79.2%), decreased smell/taste (n=89, 74.2%), and cough (n=85, 70.8%, Figure 1a). Headache, myalgias and chills were the first symptoms to peak at day 4, while cough did not peak until day 9 (Figure 1b). In 18/120 (15%) of participants, symptoms persisted through day 28 follow-up, with fatigue (7/18), cough and decreased taste/smell (5/18), and shortness of breath (4/18) reported in participants. There were significant differences in prevalence of any symptom over the entire study between participants randomized to Lambda vs placebo(8)(Figure S4A-D). Of 2730 at-home daily oral temperature measurements among 118 participants (average 23.1 measurements/participant), only 7 (0.3%) measurements in 6 participants were >100.4° F. However, oral temperatures decreased significantly after enrollment, with mean temperatures declining from 97.9°F (95% CI 97.8-98.0) between days 1-6 to 97.6°F (95% CI 97.5-97.7) between days 7-28 (coef 0.75, 95% CI 0.66-0.84, P<0.001, Figure 2a). In parallel, at home oxygen saturation measurements increased from 96.9% (95% CI 96.6-97.2) from days 1-<7 to 97.4% (95% CI 97.2-97.6) from days 7-28 (coef 0.46, 95% CI 0.21-0.72, P<0.001, Figure 2b).

**Figure 1.**
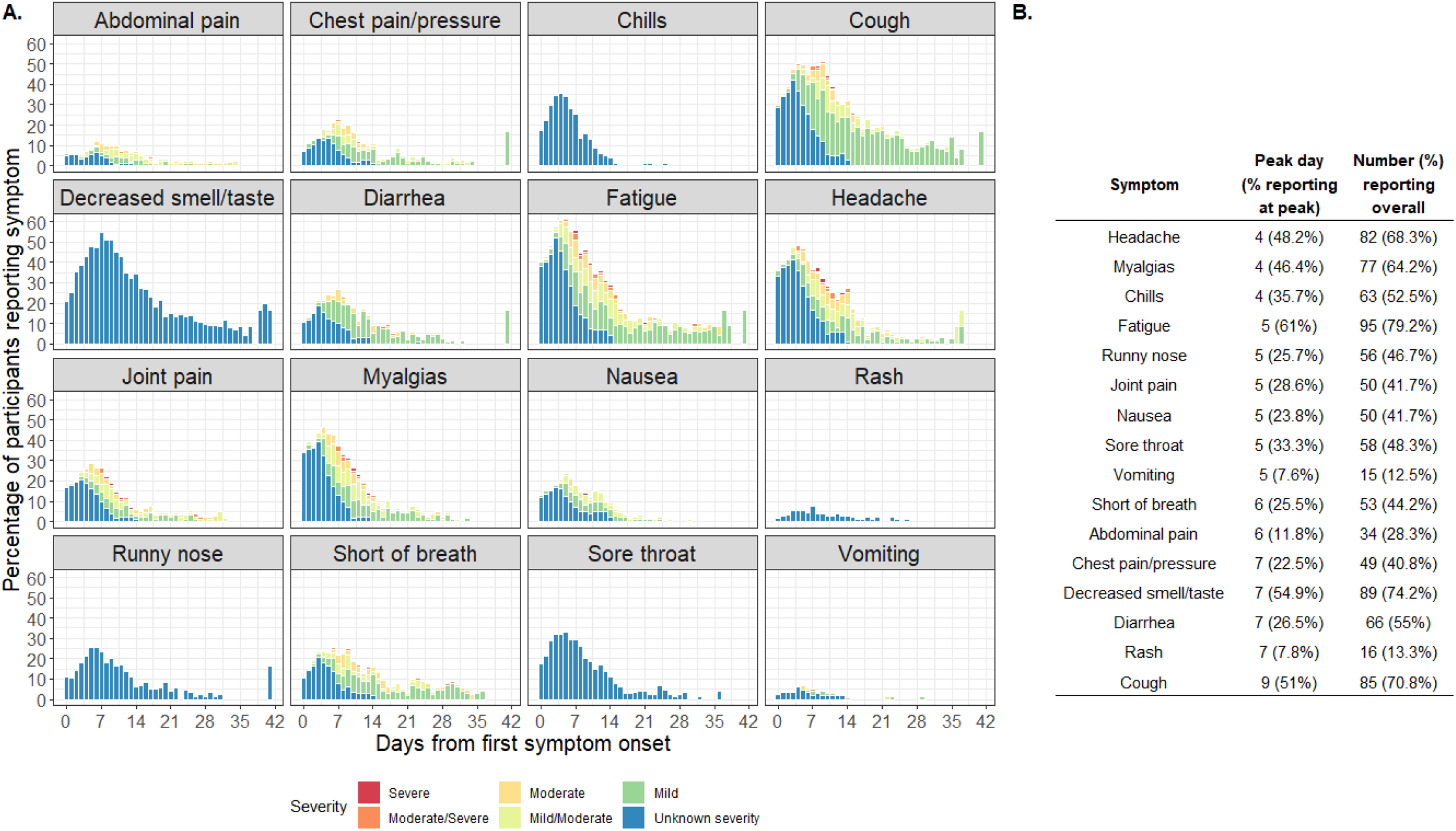
Overall Symptom Summary

**Figure 2.**
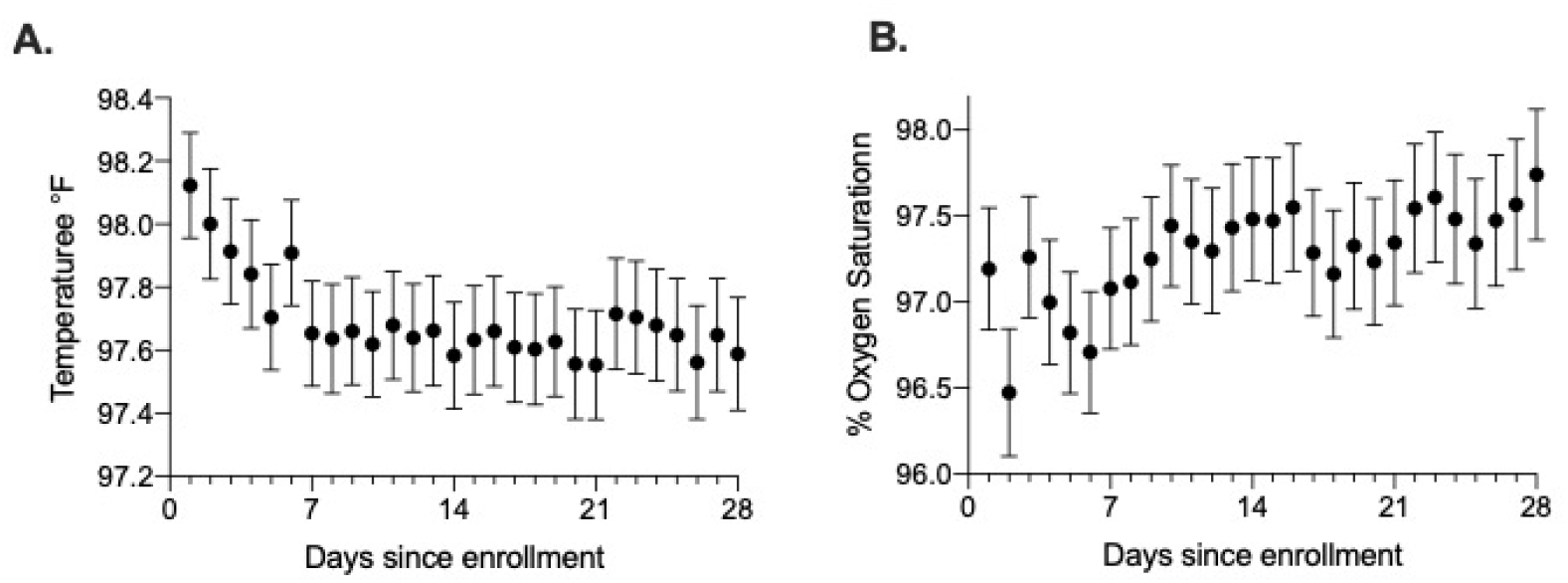
: Participant daily temperature and oxygen saturation

### Symptom Clusters

On the day of symptom onset (day 0), three symptom clusters were identified (Figure 3, Table S1, Figures S5A-F) – inflammatory (myalgias, joint pain, fatigue, headache), respiratory (shortness of breath, runny nose), and other (decreased smell/taste, chest pain/pressure), although poor within-cluster reliability was found, in which symptoms did not consistently load on the same factor (Figure 3A). The inflammatory symptom cluster persisted and increased in reliability from Day 4 to Day 10 before decreasing (Figure 3A,B). After day 0, two distinct respiratory clusters were identified, including upper (sore throat, runny nose, nausea) and lower (shortness of breath, cough, chest pain/pressure) respiratory tract symptoms. Interestingly, a larger cluster encompassing lower respiratory and inflammatory clusters emerged between days 7 and 10 of symptom onset.

**Figure 3.**
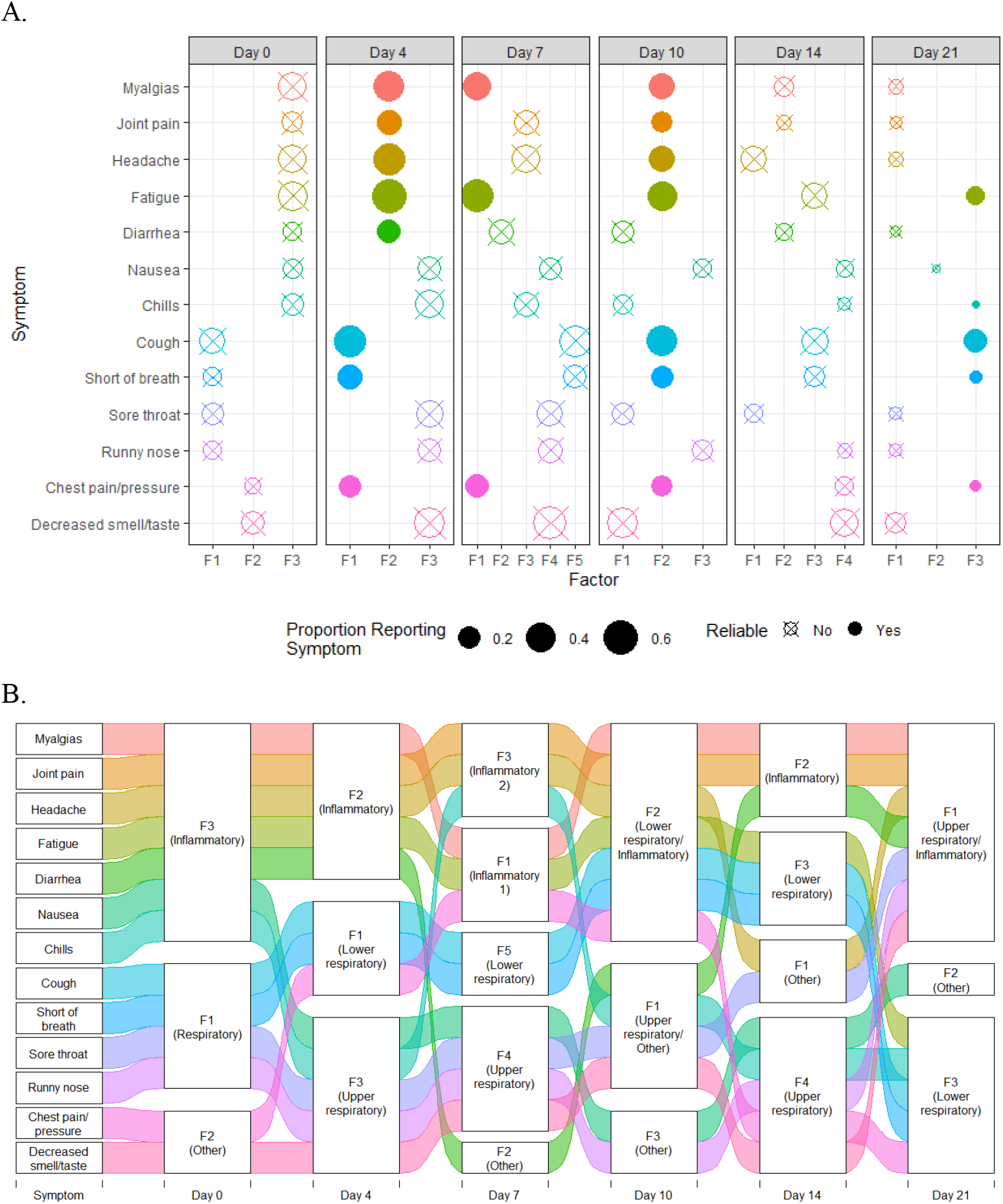
Symptom Clusters by Days from Symptom Onset as Identified by Exploratory Factor Analysis

We next assessed the joint trajectory of symptoms over time among 81 participants with symptom data, and two participant groups emerged (Figure 4). Cluster B (n=7) generally had greater symptom severity and/or later peak severity than cluster A, especially chest pain/pressure, fatigue, and myalgias. However, cluster A (n=74) had slightly higher prevalence of runny nose and sore throat compared to cluster B. There were no differences between Clusters A and B in terms of demographics, BMI, initial lab values, or baseline seropositivity (Table S2). While participants in cluster B had slightly shorter time to viral shedding cessation, these differences were not statistically significant.

**Figure 4.**
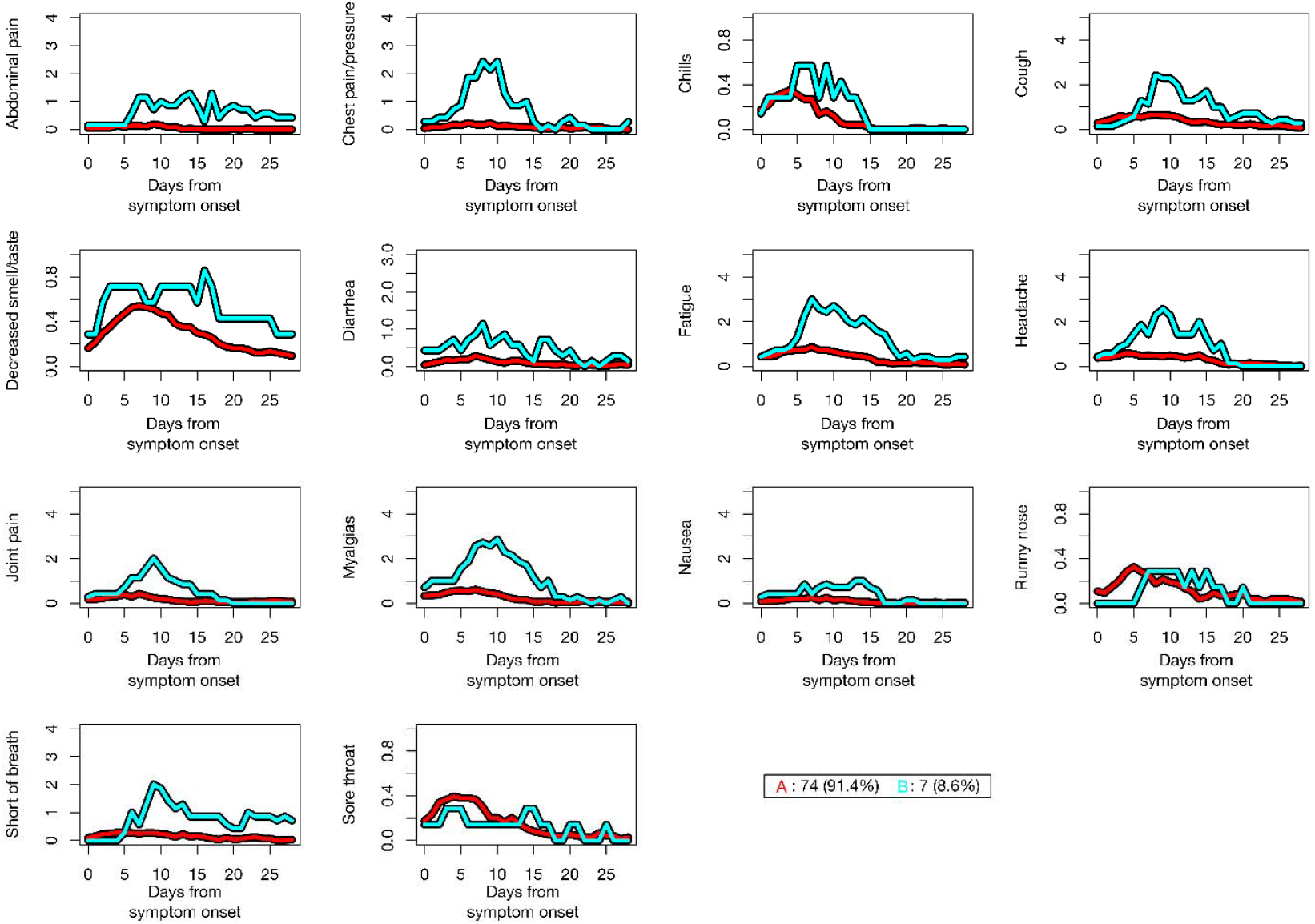
Longitudinal Clusters by Symptom

### Predictors of symptom resolution

Median time to symptom resolution from earliest symptom onset was 17 days (95% CI 14-18). Participants with higher BMI were less likely to experience symptom resolution (hazard ratio [95% CI]: 0.92 [0.85-1.0, Figure 5a). Participants identifying as LatinX had more rapid symptom resolution compared to those identifying as non-LatinX, White participants (hazard ratio [95% CI]: 2.10 [1.23-3.60]). When considering predictors of symptom resolution from the time of enrollment, for every 10 unit increase in ALT at enrollment, likelihood of symptom resolution increased by 10% (Figure 5b). Neither enrollment antibody levels nor oropharyngeal swab viral loads were significantly associated with symptom resolution (Figure 5b). Results were similar when stratifying by treatment arm (data not shown).

**Figure 5.**
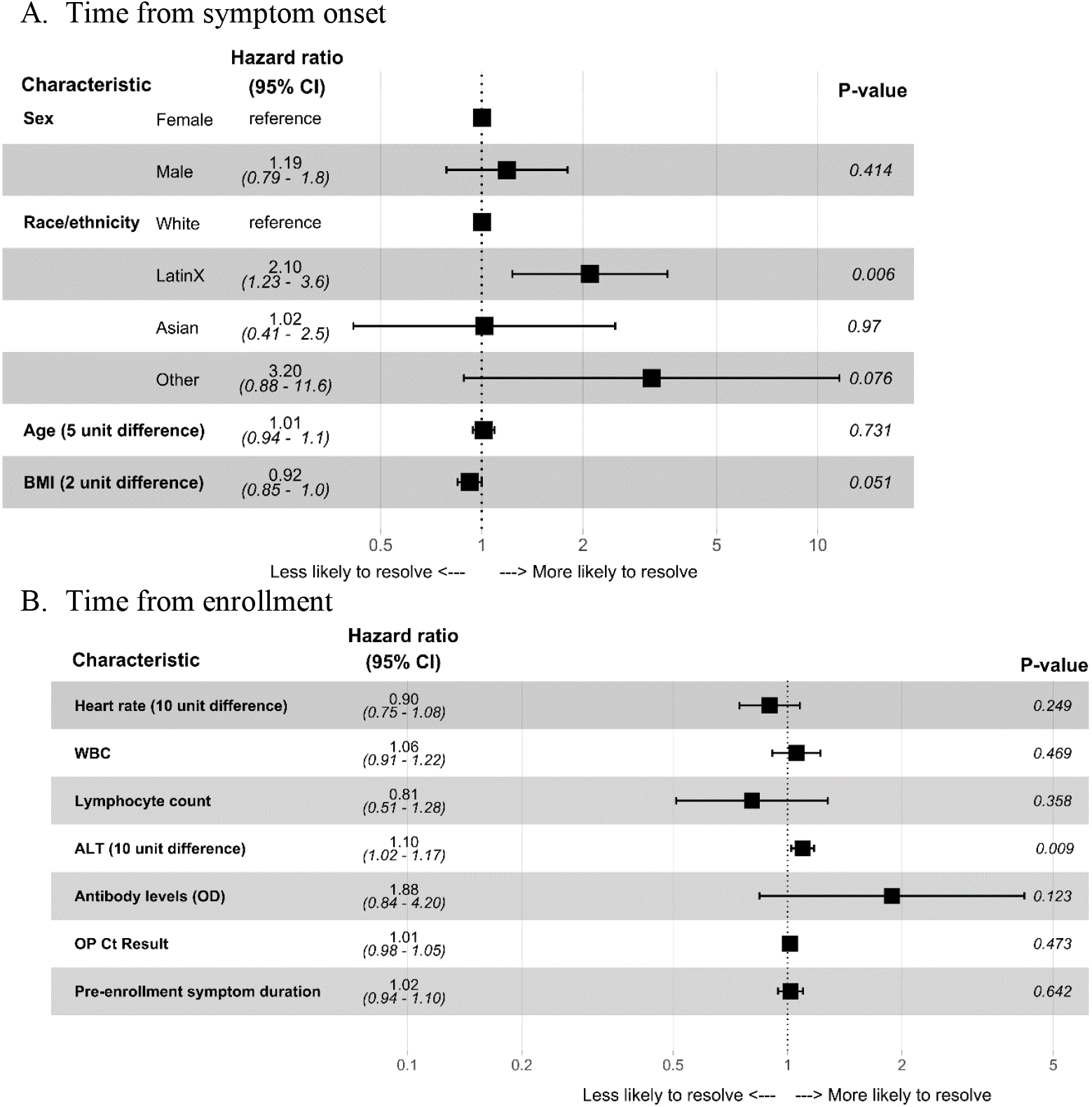
Hazard Ratios for Time to Symptom Resolution

### Virologic trajectories and predictors of viral shedding cessation

The median time to cessation of oropharyngeal viral shedding was 10 days from first reported positive test (95% CI: 8-12), with 108 participants experiencing cessation of viral shedding during the study. Higher SARS-CoV-2 IgG levels (HR [95% CI]: 3.3 [1.5, 7.2]) at enrollment were associated with more rapid viral shedding cessation (Figure 6). Higher cycle threshold (Ct) values at enrollment were also associated with more rapid viral shedding cessation in an independent model (HR [95% CI]: 1.2 [1.1, 1.2], p<0.001). No other baseline demographics or measured laboratory values were significantly associated with viral shedding cessation (Figure 6). Results were similar when stratifying by treatment arm (data not shown).

**Figure 6.**
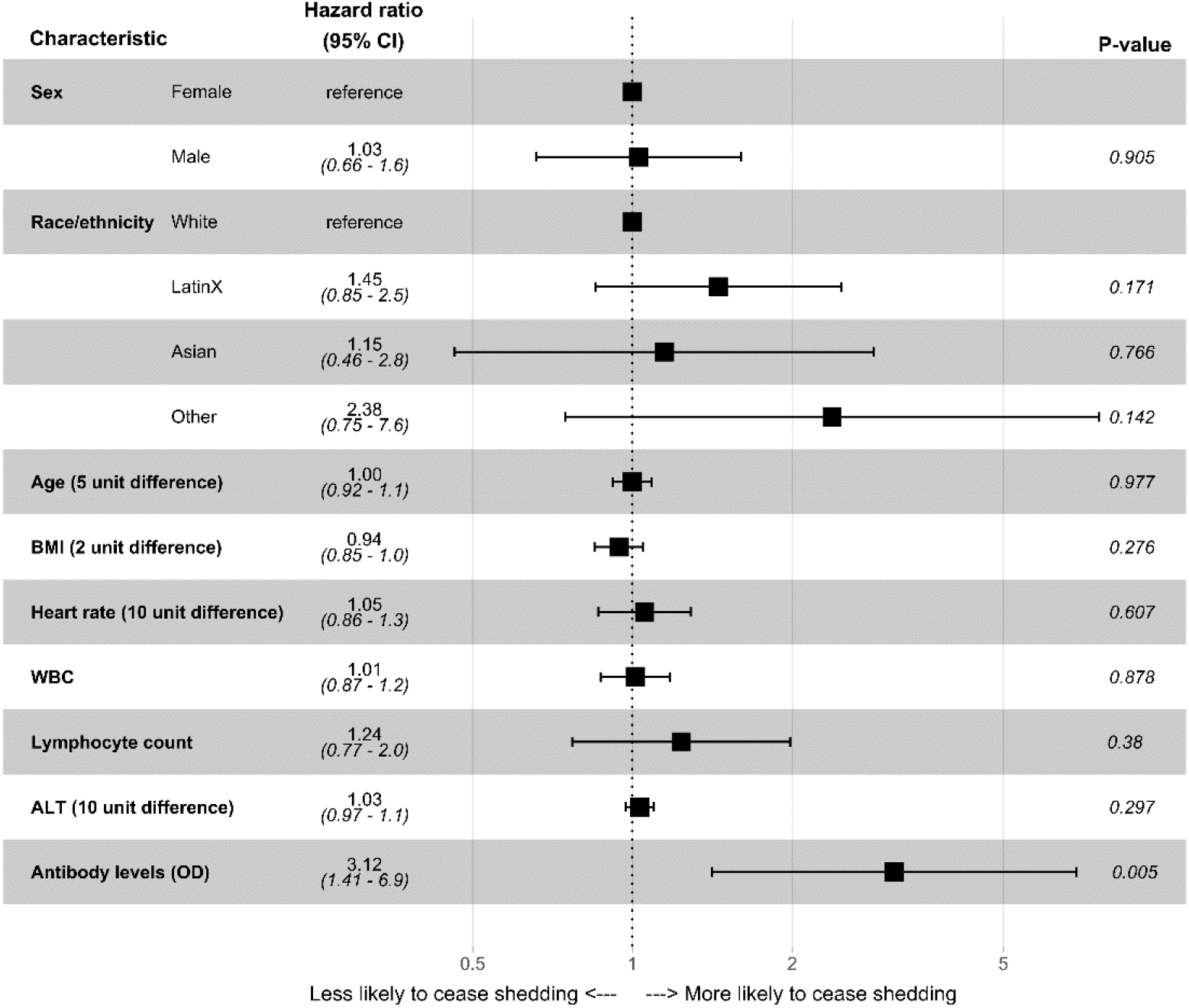
Hazard Ratios for Time to Shedding Cessation

### Associations between clinical symptoms and oropharyngeal viral shedding

We visually assessed antibody levels over time (Figure 7A) and whether time from enrollment until viral shedding cessation and symptom resolution were similar in our cohort (Figure 7B). At enrollment, 40.8% (49/120) study participants were SARS-CoV-2 seropositive; this increased to 87.7% (100/114) by day 5, and 95.5% (106/111) by day 14 (Figure 7A). Half of participants included stopped shedding virus by 7 days after enrollment, while half of participants experienced symptom resolution 2 days later on study day 9. While most participants experienced shedding cessation before their symptoms resolved, the survival probability was similar over time for both outcomes.

**Figure 7.**
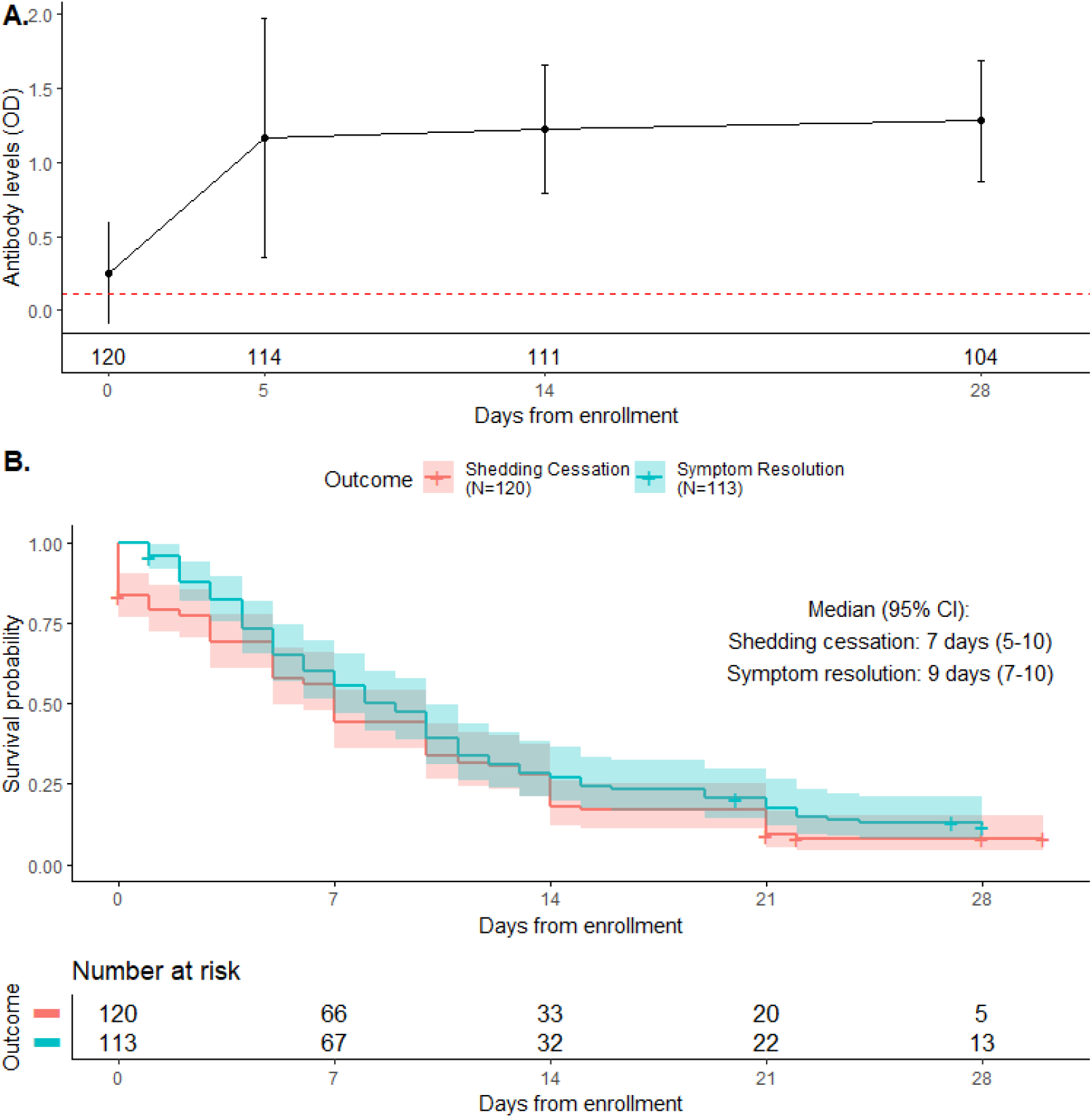
Antibody levels over time and joint KM curves of time to shedding cessation and time to symptom resolution

Finally, we assessed whether specific symptoms were associated with detection of oropharyngeal SARS-CoV-2 RNA. On each day, the majority of participants reporting symptoms had detectable oropharyngeal SARS-CoV-2 RNA (Figure 8A). In multivariable analysis, joint pain (OR 2.88, 95% CI 1.39-5.99), myalgias (OR 2.71, 95% CI 1.51-4.84), chills (OR 2.4, 1.16-4.99), and fatigue (OR 1.68, 95% CI 1.01-2.81) were associated with significantly greater odds of oropharyngeal SARS-CoV-2 RNA detection (Figure 8B). In contrast, there was no statistically significant association between presence or absence of upper respiratory symptoms (e.g., runny nose, shortness of breath, sore throat, anosmia) and odds of oropharyngeal SARS-CoV-2 RNA detection.

**Figure 8.**
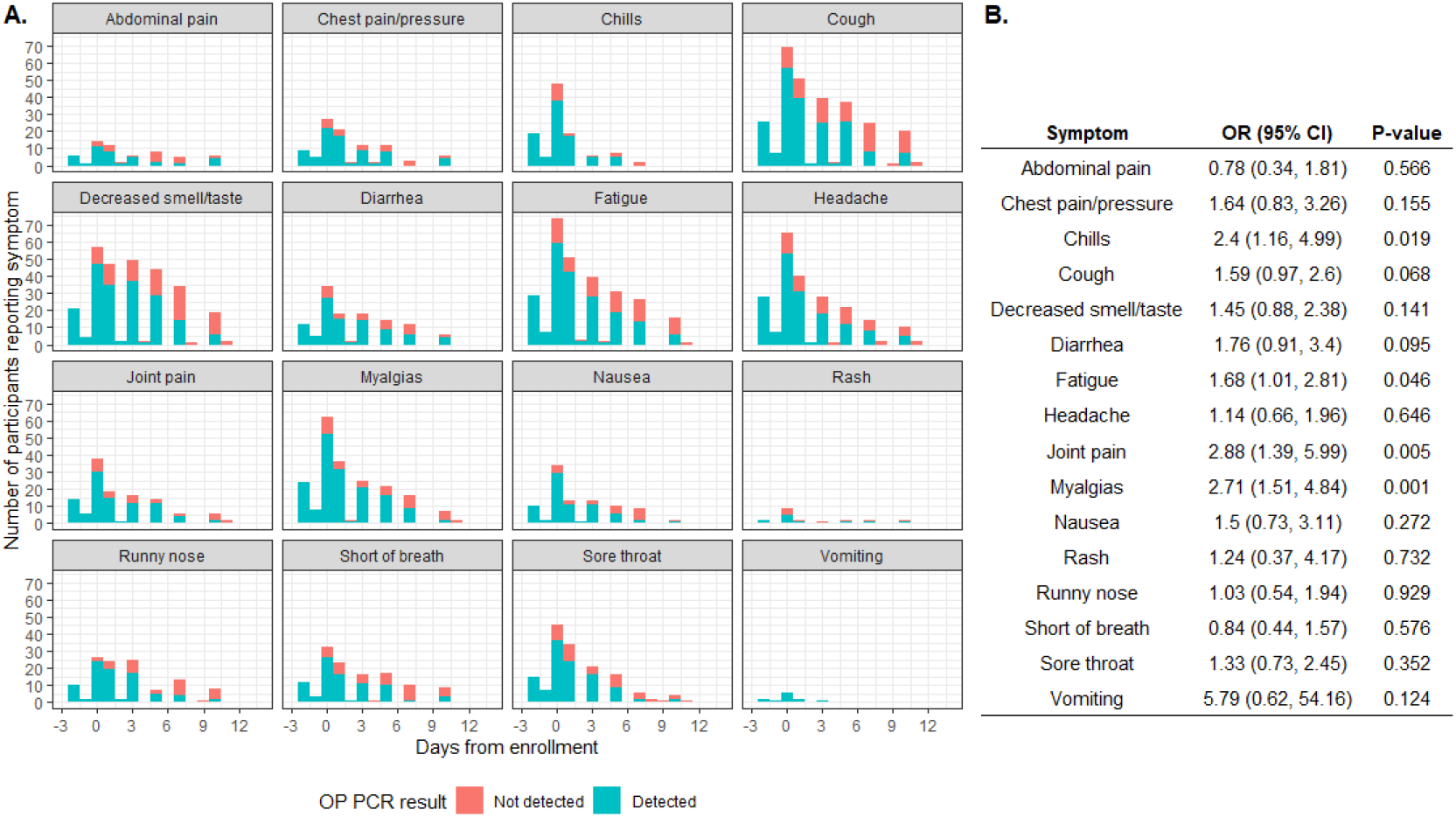
OP PCR Status by Reported Symptoms Over Time

## Discussion

In this study, we leveraged a comprehensive and detailed dataset among outpatients with uncomplicated COVID-19 enrolled in a Phase 2 clinical trial to carefully define symptom prevalence, longitudinal emergence and resolution of individual symptoms and symptom clusters, and associations between symptoms, viral shedding, and SARS-CoV-2 antibody levels. Inflammatory symptoms (myalgias, chills, fatigue) were most prevalent and peaked early; these symptoms were also associated with ongoing viral replication. In contrast, objective fever and respiratory symptoms were less common, and not associated with viral replication. Though higher SARS-CoV-2 IgG levels at enrollment were associated with more rapid viral shedding cessation, they were not associated with time to symptom resolution. Participants were enrolled early in the course of disease, with excellent participant retention and little missing data. Only 4 participants were hospitalized, and no deaths observed.

Importantly, though the most prevalent symptoms observed were consistent with previous reports,(3, 4, 6, 11, 12) we observed different prevalence peaks per symptom. The most common initial symptoms were headache, myalgias and chills; these symptoms peaked at 4 days post symptom onset. Shortness of breath and cough peaked at 6 and 9 days, respectively. These data differ from a prediction model which suggested that cough would precede headache and myalgias.(13) Anosmia was common and is likely specific for COVID-19(14), but was most prominent one week post symptom onset. Thus, patients may be shedding infectious virus for up to one week prior to the onset of “typical” COVID-19 symptoms such as cough, shortness of breath, and anosmia.

Only 3 participants had fever >100.4° F at enrollment, and 6 had fever at any timepoint during follow-up, consistent with other studies which show relative lack of fever in mild/moderate cases compared with severe/critical cases.(3-7) However, overall temperature did decrease in the first week after enrollment, after which temperatures remained stable at a mean temperature of 97.6°F. These observations are consistent with recent reports suggesting that “normal” body temperatures are well below the established “normal” of 98.6°F,(15, 16) and suggest that temperatures above 97.6°F may represent clinically significant physiologic changes within the context of uncomplicated COVID-19.

LatinX participants were significantly more likely to have symptoms resolve at any point during the study compared to non-LatinX, White participants. This was after controlling for key demographic/clinical characteristics, including age, gender, and BMI. Although it is possible that biologic differences may underlie these differences in symptom resolution, prior reports have suggested that race or ethnic-based differences are not significant predictors of clinically relevant hospitalization outcomes such as mortality, ICU admission, or mechanical ventilation.(17, 18) These differences may be driven by residual confounding, including the possibility of differential symptom recall bias and/or delays in testing/diagnosis prior to enrollment.

The median time to viral shedding cessation from enrollment was 10 days, consistent with other reports.(19, 20) Though other studies have identified disease severity as a risk factor for longer duration of shedding(20), in this cohort, only SARS-CoV-2 antibody levels and oropharyngeal viral loads at enrollment were associated with more rapid viral shedding cessation. The majority of participants reporting symptoms on each day had detectable oropharyngeal virus by PCR, suggesting that ongoing presence of symptoms is an indicator of ongoing viral shedding. This supports CDC guidelines(21) to continue isolating until symptoms are improved, though PCR positivity does not infer infectivity.(22) Myalgia and joint pain were the only symptoms significantly associated with oropharyngeal PCR positivity. These are also some of the earliest symptoms to appear, when viral loads are higher early in disease course and patients are most likely to be infectious. Therefore, screening for respiratory symptoms may miss the most infectious patients, who may attribute their symptoms to other causes.

There were limitations to this study. This cohort was part of a randomized, placebo-controlled trial, where half the cohort received Lambda and the other half received placebo. The duration of symptoms and viral shedding was nearly identical between the two arms in the primary analysis(8), so we included the entire cohort in order to improve statistical power to perform our analyses. Only 4 (3%) participants in this cohort needed hospitalization and 10 (8.3%) went to the ED for evaluation, compared with an incidence of hospitalization of 2.9-11% in other outpatient trials.(7, 23) This is likely related to study selection criteria, and that recruitment occurred during non-surge conditions when even moderately ill patients were being hospitalized. Median participant age was 36 years, and the cohort was predominantly LatinX, reflecting our local population of COVID-19 cases(24); however, our findings may not be generalizable to other settings. Though there was high retention in the study, 34 participants missed two consecutive daily symptom surveys and were excluded from the symptom cluster analysis. Though we collected symptom data daily for 28 days after enrollment, time of symptom onset was reported retrospectively by participants at enrollment and thus subject to recall bias. Finally, this is a secondary analysis of data from a clinical trial that was not designed for these analyses, so this study should be considered hypothesis-generating.

In this cohort of participants with uncomplicated COVID-19, distinct symptom clusters and trajectories were identified and inflammatory symptoms were associated with ongoing viral replication. In contrast, cough, loss of taste/smell, and elevated temperature, which are often used for screening programs, are not always earliest to present and not associated with viral replication, and may therefore not be ideal for screening approaches. Data from this and other longitudinal studies will be critical in shaping our understanding of disease pathogenesis in patients, and for planning large, Phase 3 outpatient studies based on clinical and virologic endpoints.

## Data Availability

Data from this clinical trial is available in the supplemental materials of the parent study publication.

## Acknowledgements

We would like to thank the Lambda study participants, the entire study team and the Clinical and Translational Research Unit staff. We are grateful to Catherine Blish for her work establishing the Biobank and Taia Wang for her work on the COVID-19 serologies.

---

Figure 1. A) Distribution of the percentage of participants reporting each symptom by day from symptom onset. Color represents the severity of the symptom reported. ‘Unknown severity’ includes both symptoms in which severity was not asked and symptoms reported as starting prior to enrollment. B) Summary of the peak day of which each symptom was reported with corresponding percentage (%) of participants reporting the symptom on the peak day, and the number and percentage of participants reporting each symptom at least once during the study period. For symptoms present prior to enrollment, severity data was not collected.

Figure 2. Temperature (A) and oxygen saturation (B) was measured daily at home using study-provided equipment. Shown are mean temperature and mean percent oxygen saturation with 95% confidence intervals (marginal estimates from GEE models) on each day after enrollment.

Figure 3: Results from the exploratory factor analysis of symptoms. Results are colored by symptom. A) Dot plot of symptoms by days from symptom onset and resulting factors. Size of dot represents the proportion of participants reporting that symptom on that day. Symptoms that load on a factor are represented by a symbol (reliable vs not). Within-factor reliability is defined as a Cronbach’s alpha of 0.70 and above. B) Alluvial plot of how symptoms group into clusters/factors and how those groups change over time.

Figure 4: Joint cluster trajectories by derived cluster and symptom. Y-axis represents the severity score of the symptom (0-5 or 0-1) while the x-axis represents days from symptom onset.

Figure 5: Forest plots of the multivariable Cox proportional hazards model for A) time from symptom onset and B) time from enrollment to first day of symptom resolution. Vital signs and clinical labs were measured at enrollment. Time from enrollment model additionally adjusted for sex, age, race/ethnicity, and BMI. Visualizations of each hazard ratio are presented on the log-HR scale. WBC = white blood count, ALT = alanine transaminase, OD = optical density, OP=oropharyngeal, Ct=cycle threshold. N=111 (# events=96) and N=107 (# events=93), respectively.

Figure 6: Forest plot of the multivariable Cox proportional hazards model for time from first positive PCR test to time to first of two consecutive negative PCR tests. Vital signs and clinical labs were measured at enrollment. Visualizations of each hazard ratio are displayed on the log-HR scale. WBC = white blood count, ALT = alanine transaminase, OD = optical density, OP=oropharyngeal. N=114, # events = 102.

Figure 7: A) Mean antibody levels measured over the study period. Bars represent the standard deviation around the mean and count below each bar represents the number of participants contributing data at that study day. Red dashed line represents the threshold of seropositivity, where values above the dashed line indicate seropositive. B) Joint Kaplan-Meier survival plots of time to shedding cessation and symptom resolution from enrollment. Lines are not mutually exclusive. OD = optical density.

Figure 8: A) Stacked bar plots of number of participants reporting symptoms on each day by symptom and oropharyngeal (OP) PCR result. Within each bar, color represents the proportion of those participants with either PCR detected or not detected. B) mixed effect logistic regression model results for odds of a positive PCR as a function of symptom reported (yes/no), age at enrollment, sex, treatment arm, and time from first positive result. Odds ratios (ORs) in each row correspond to a separate symptom model.

