## Supplementary Materials for "Inflammatory, but not respiratory symptoms, associated with ongoing upper airway viral replication in outpatients with uncomplicated COVID-19"

**Clinical Trials Registration**: NCT04331899

**Supplementary Methods**

**Laboratory procedures**

Laboratory measurements were performed by trained study personnel using point-of-care CLIA-waived devices or in the Stanford Health Care Clinical Laboratory. Oropharyngeal swabs were tested for SARS-CoV-2 in the Stanford Clinical Virology Laboratory using an emergency use authorized, laboratory-developed, RT-PCR.(1, 2) (3) Centers for Disease Control and Prevention guidelines identify oropharyngeal swabs as acceptable upper respiratory specimens to test for the presence of SARS-CoV-2 RNA,(4) and detection of SARS-CoV-2 RNA swabs using oropharyngeal swabs was analytically validated in the Stanford virology laboratory. A positive RT-PCR test was defined by a cycle threshold (Ct) value <42 for both N1 and N3 gene regions. A negative test was defined as failure to detect SARS-CoV-2 viral RNA by 42 cycles for either gene regions.

IgG antibody levels against the SARS-CoV-2 spike receptor binding domain (RBD) were assessed at enrollment and day 5, 14, and 28 post-enrollment.(5) Briefly, heat inactivated serum samples at enrolment were diluted 5-fold starting at 1:50 and IgG antibody levels against RBD determined by ELISA. Absorbance was measured at 450nm (SPECTRAmax 250, Molecular Devices). Samples were considered seropositive against RBD if their absorbance value was greater than the mean plus four standard deviation (SD) of all negative controls (n=130).

**Statistical Analysis:**

*Symptom Cluster Analysis*

Symptoms with more than 5% overall study prevalence were chosen for examination in exploratory factor analysis (EFA), since symptoms with lower prevalence were most likely not key symptoms, thus would be unstable for cluster identification. The maximum likelihood procedure was applied in each EFA using an oblique rotation to allow factors to be correlated.(6) Parallel analysis, scree plots, and the minimum average partial (MAP) procedures were used to determine the appropriate number of clusters on each of the 6 days of interest.(7) The reliability of derived factors was assessed by Cronbach’s alpha.(8) Changes in factors derived and symptoms contributing to each factor were compared descriptively across time points of interest.

The joint trajectory of symptoms was also assessed to determine whether participant clusters persist over time. Participants with more than two consecutive days of missing symptom data were excluded. Among the remaining participants, missing values were imputed using a last observation carried forward approach assuming symptom presence/absence at the last reported day remained unchanged. The method proposed by Genolini et al. was applied to symptom data recorded in the first 28 days after symptom onset.(9) This method implements a *k*-means approach designed specifically for joint trajectories by choosing the number of clusters using an iterative approach, and attributing participants to clusters such that individuals in the same cluster have similar symptom trajectories. The quality criteria proposed by Calinski and Harabasz criterion was used to assess within-cluster and between-cluster indices.(10) Fisher’s exact test and the Kruskal-Wallis rank sum test were used to test for differences in baseline characteristics and demographics by cluster assignment.

SUPPLEMENTAL TABLES

Table S1. Factor Loadings by Days from Symptom Onset

| **Symptom** | **Day 0** | | | **Day 4** | | | **Day 7** | | | | | **Day 10** | | | **Day 14** | | | | **Day 21** | | |
| --- | --- | --- | --- | --- | --- | --- | --- | --- | --- | --- | --- | --- | --- | --- | --- | --- | --- | --- | --- | --- | --- |
|  | **F1** | **F2** | **F3** | **F1** | **F2** | **F3** | **F1** | **F2** | **F3** | **F4** | **F5** | **F1** | **F2** | **F3** | **F1** | **F2** | **F3** | **F4** | **F1** | **F2** | **F3** |
| Chest pain/pressure |  | 0.39 |  | 0.39 |  |  | 0.49 |  |  |  | 0.37 |  | 0.81 |  |  |  |  | 0.79 |  |  | 0.95 |
| Chills |  |  | 0.38 |  |  | 0.33 |  |  |  |  |  | 1.00 |  |  |  |  |  | 0.36 |  |  | 0.99 |
| Cough |  |  |  | 0.47 |  |  |  |  |  |  | 0.48 |  | 0.64 |  |  |  | 0.55 |  |  |  | 0.39 |
| Decreased smell/taste |  | 0.84 |  |  |  | 0.56 |  |  |  |  |  |  |  |  |  |  |  | 0.35 |  |  |  |
| Diarrhea |  |  |  |  |  |  |  | 1.01 |  |  |  | 0.46 |  |  |  | 0.4 |  |  |  |  |  |
| Fatigue |  |  | 0.43 |  | 0.45 |  | 1.01 |  |  |  |  |  | 0.74 |  |  |  | 1.00 |  |  |  | 0.74 |
| Headache |  |  | 0.38 |  | 0.73 |  |  |  | 0.44 |  |  |  | 0.64 |  | 0.42 | 0.37 |  |  | 0.85 |  |  |
| Joint pain |  |  | 0.51 |  | 0.79 |  |  |  | 0.94 |  |  |  | 0.60 |  |  | 0.47 |  |  | 0.33 |  |  |
| Myalgias |  |  | 0.75 |  | 0.69 |  | 0.52 |  | 0.43 |  |  |  | 0.76 |  |  | 0.98 |  |  |  |  |  |
| Nausea |  |  | 0.46 |  |  | 0.48 |  |  |  | 0.37 |  |  |  |  |  |  |  | 0.51 |  | 0.99 |  |
| Runny nose | 0.36 |  |  |  |  |  |  |  |  | 0.49 |  |  |  | 0.87 |  |  |  | 0.39 | 0.41 |  |  |
| Short of breath | 0.90 |  |  | 1.02 |  |  |  |  |  |  | 0.45 |  | 0.76 | 0 |  |  | 0.31 |  |  |  | 0.91 |
| Sore throat |  |  |  |  |  |  |  |  |  | 0.70 |  | 0.42 |  | 0.36 | 0.97 |  |  |  | 0.89 |  |  |
| Cronbach’s alpha | 0.41 | 0.46 | 0.63 | 0.71 | 0.73 | 0.38 | 0.75 | NA | 0.60 | 0.44 | 0.52 | 0.61 | 0.87 | 0.42 | 0.46 | 0.69 | 0.68 | 0.66 | 0.40 | NA | 0.81 |

Note: only loadings > 0.30 are reported.

Table S2. Baseline Characteristics by Longitudinal Cluster

| Characteristic | Cluster | | P-value |
| --- | --- | --- | --- |
|  | A  (n=74) | B  (n=7) |  |
| Male, n (%) | 44 (59.5) | 5 (71.4) | 0.70 |
| Age, mean (SD) | 39.31 (13.36) | 45.86 (17.62) | 0.23 |
| Race/ethnicity, n (%) |  |  |  |
| LatinX | 41 (55.4) | 5 (71.4) | 0.85 |
| White | 26 (35.1) | 2 (28.6) |  |
| Asian | 6 ( 8.1) | 0 ( 0.0) |  |
| Other | 1 ( 1.4) | 0 ( 0.0) |  |
| BMI, mean (SD) | 28.61 (5.76) | 29.05 (5.05) | 0.85 |
| Heart rate, mean (SD) | 78.51 (13.37) | 84.00 (15.82) | 0.31 |
| Oxygen saturation, median [IQR] | 99.00 [97.00, 99.00] | 97.00 [96.50, 97.50] | 0.047 |
| WBC, median [IQR] | 5.00 [3.92, 6.45] | 4.70 [3.40, 5.55] | 0.33 |
| Lymphocyte count, median [IQR] | 1.41 [1.15, 1.88] | 1.20 [0.91, 1.95] | 0.37 |
| ALT, median [IQR] | 30.50 [21.25, 48.75] | 38.00 [27.00, 45.00] | 0.71 |
| AST, median [IQR] | 30.00 [25.00, 39.75] | 39.00 [34.50, 42.00] | 0.07 |
| Antibody level (OD), median [IQR] | 0.01 [-0.02, 0.43] | 0.04 [0.00, 0.16] | 0.69 |
| Seropositive, n (%) | 24 (32.4) | 2 (28.6) | 1.00 |
| PCR Not detected, n (%) | 14 (19.2) | 1 (14.3) | 1.00 |
| Viral load, median [IQR] | 6538.25 [355.75, 191153.06] | 10609.65 [4094.20, 57471.53] | 0.89 |

WBC = white blood count, ALT = alanine transaminase, OD = optical density.

Table S3. Univariate associations with seropositivity at Day 28

|  | Seronegative  (n=5) | Seropositive  (n=57) | P-value |
| --- | --- | --- | --- |
| Male, n (%) | 2 (40.0) | 37 (64.9) | 0.35 |
| Age, mean (SD) | 40.00 (14.20) | 37.54 (12.78) | 0.68 |
| Race/ethnicity, n (%) |  |  |  |
| LatinX | 1 (20.0) | 30 (52.6) | 0.14 |
| White | 3 (60.0) | 21 (36.8) |  |
| Asian | 0 ( 0.0) | 5 ( 8.8) |  |
| Other | 1 (20.0) | 1 ( 1.8) |  |
| BMI, mean (SD) | 24.23 (7.25) | 29.02 (5.68) | 0.08 |
| Heart rate, mean (SD) | 81.20 (21.68) | 80.45 (12.60) | 0.91 |
| Oxygen saturation, median [IQR] | 100.00 [98.00, 100.00] | 98.00 [97.00, 99.00] | 0.38 |
| WBC, median [IQR] | 8.00 [7.90, 8.50] | 4.40 [3.60, 5.50] | 0.001 |
| Lymphocyte count, median [IQR] | 2.40 [1.83, 2.60] | 1.35 [1.10, 1.70] | 0.006 |
| ALT, median [IQR] | 26.00 [21.00, 30.00] | 34.00 [22.00, 49.00] | 0.41 |
| AST, median [IQR] | 29.00 [25.00, 29.00] | 30.00 [26.00, 41.00] | 0.28 |
| Antibody level (OD), median [IQR] | -0.02 [-0.02, -0.01] | -0.01 [-0.02, 0.02] | 0.26 |

WBC = white blood count, ALT = alanine transaminase, OD = optical density.

SUPPLEMENTAL FIGURES

Figure S1: Lambda trial CONSORT diagram

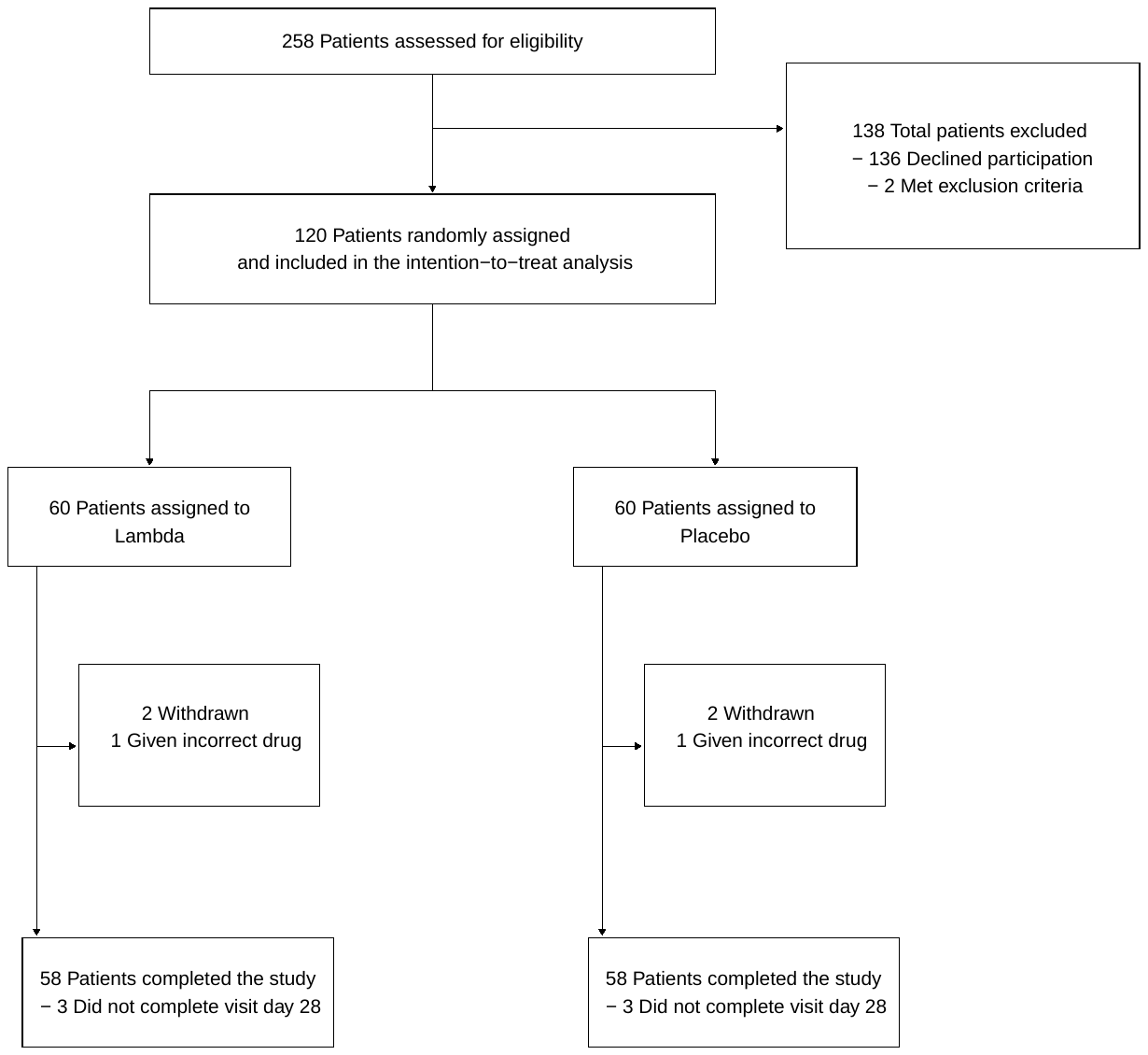

Figure S2: Antibody level by days since onset of symptoms

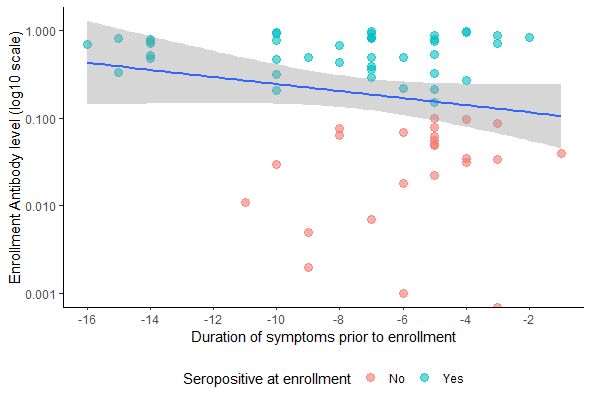

Figure S2: Dot plot of enrollment antibody levels by duration of symptoms prior to enrollment and seropositivity at enrollment. Blue line represents the slope based on a simple linear regression. N=107.

Figure S3: Symptom Summaries

A. Respiratory Symptom Summary

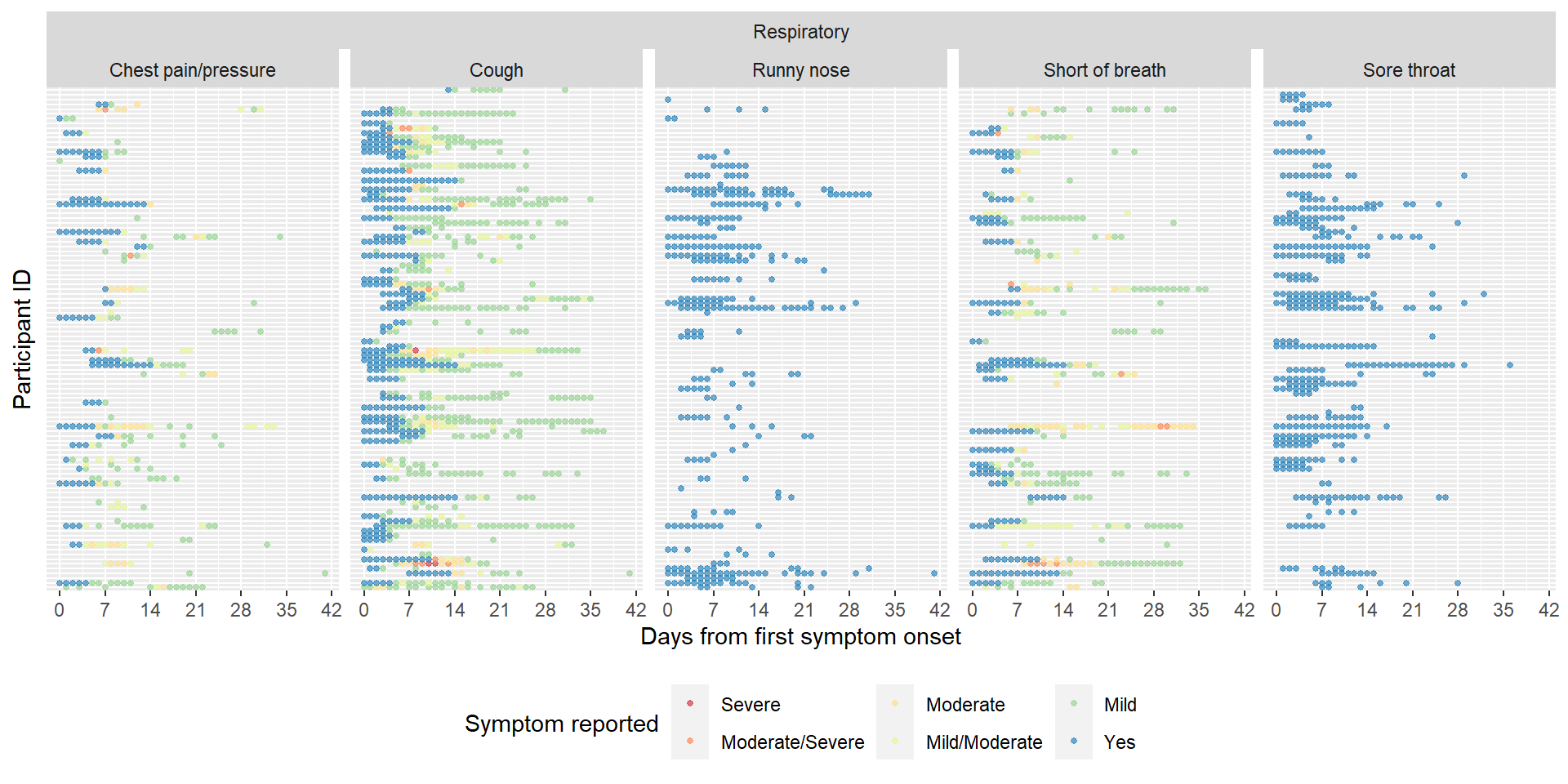

B. Systemic Symptom Summary

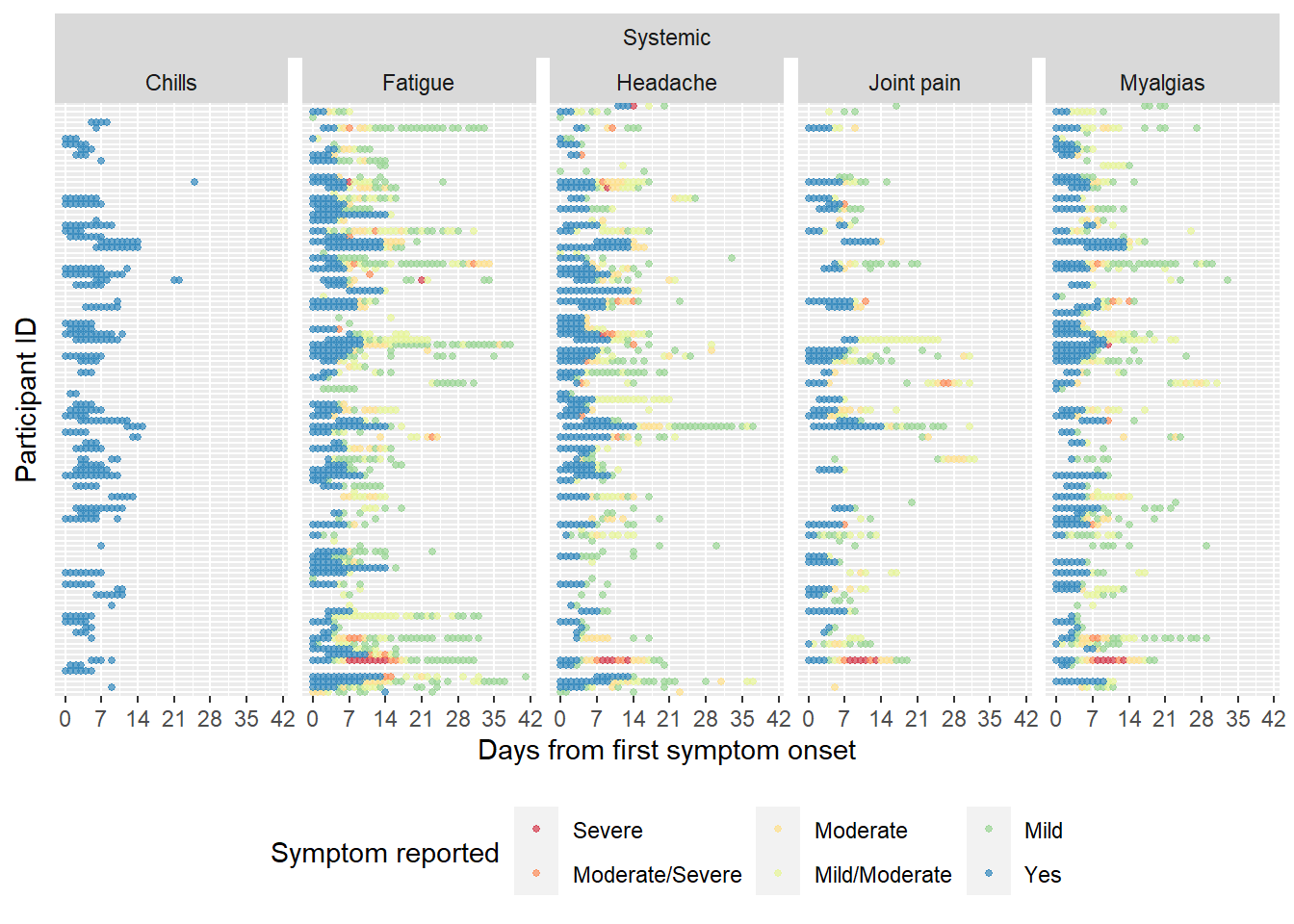

C. GI Symptom Summary

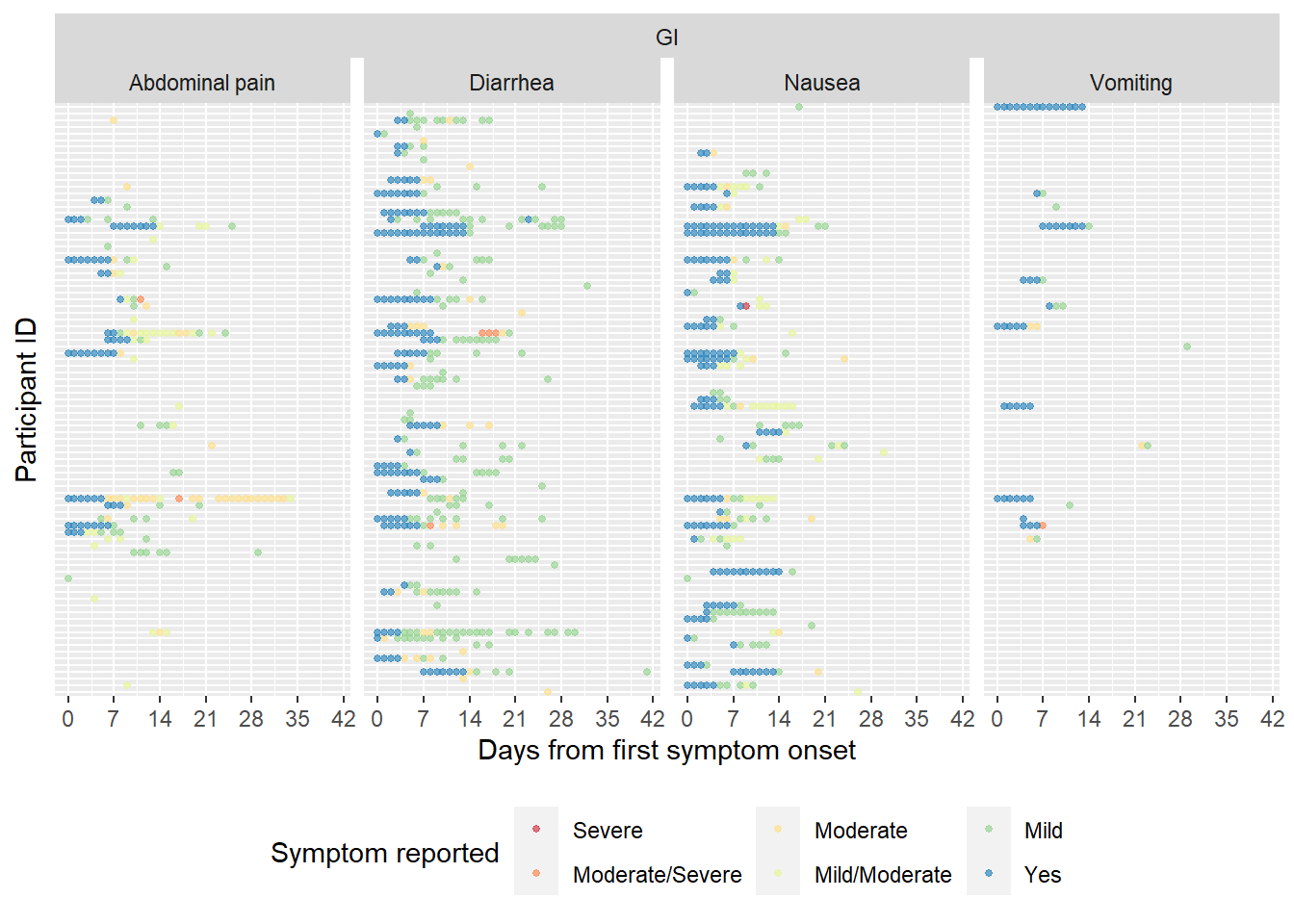

D. Other Symptom Summary

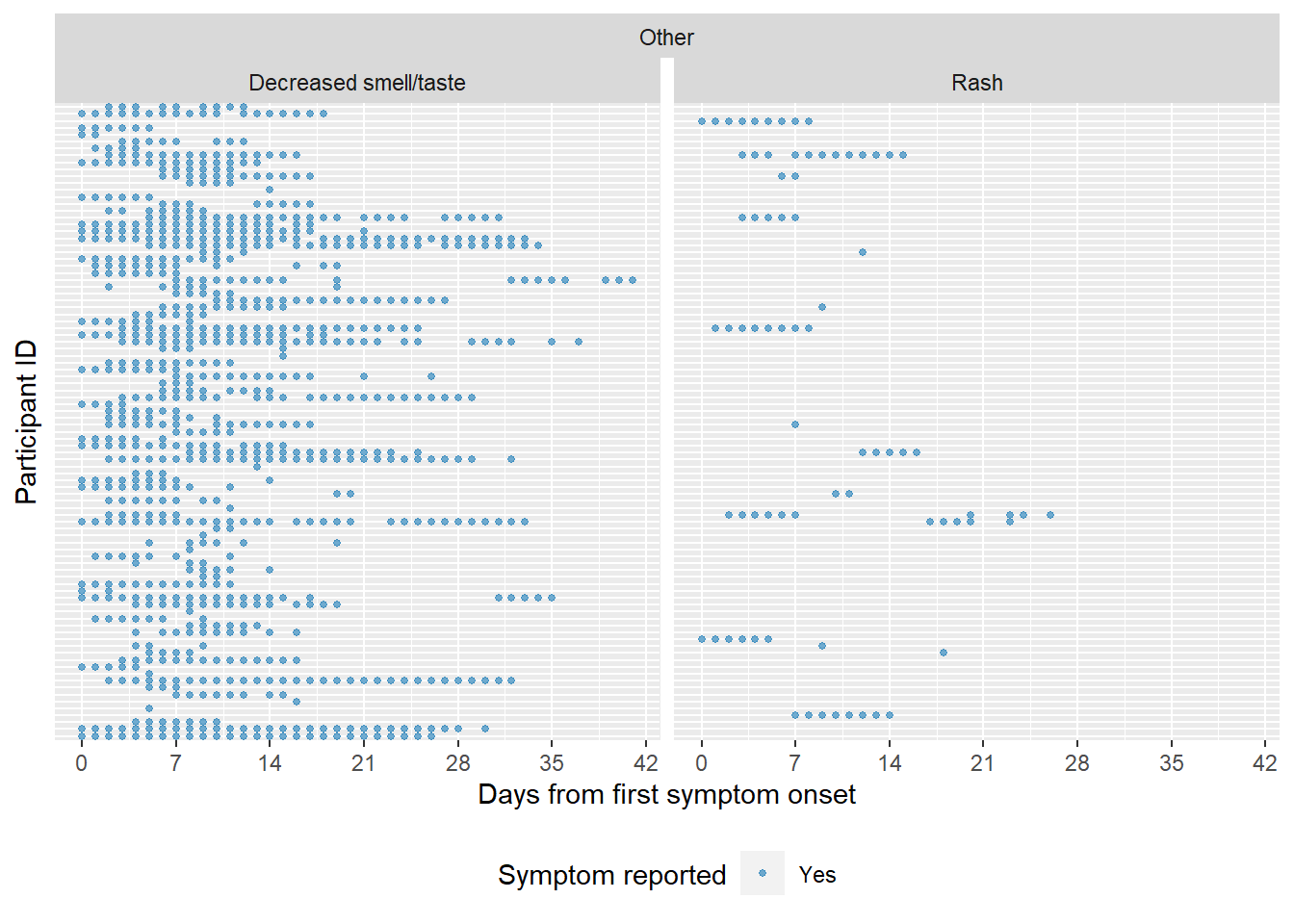

Figure S4. Distribution of Symptoms by Treatment Arm

A. Respiratory

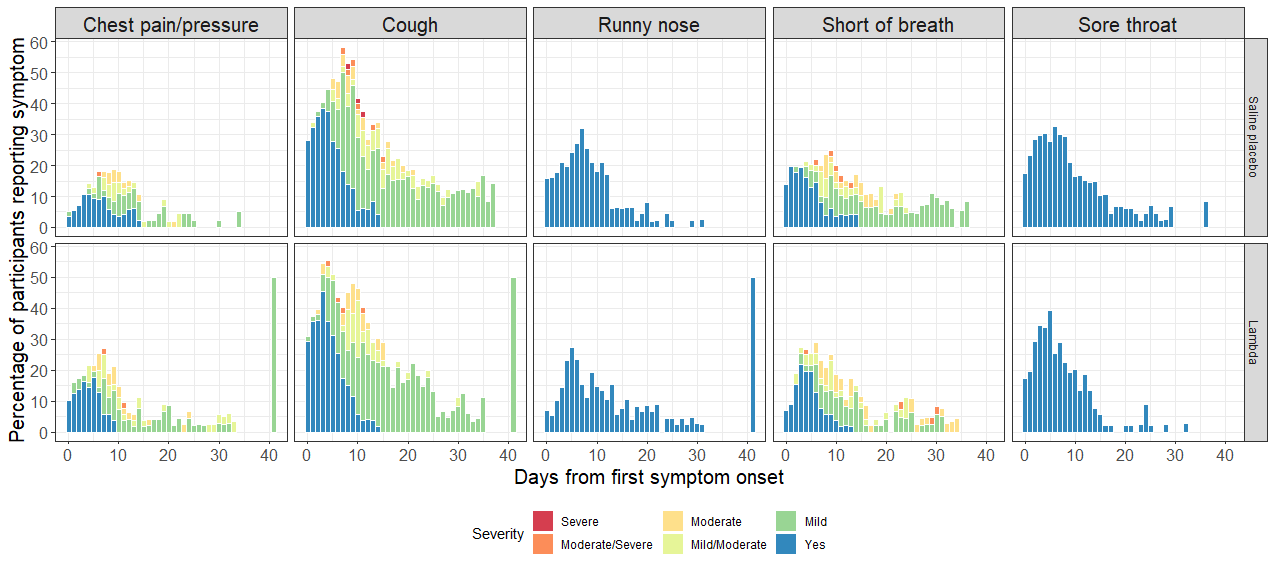

B. Systemic

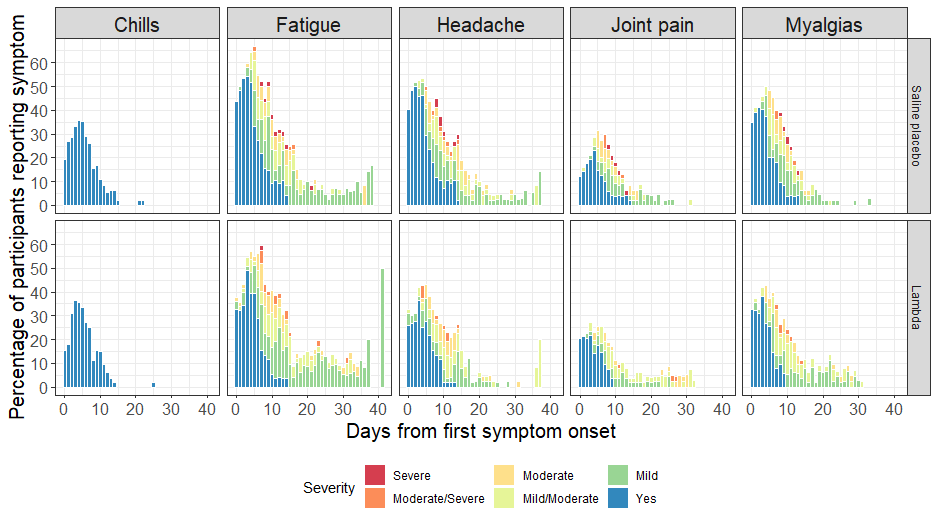

C. Gastrointestinal

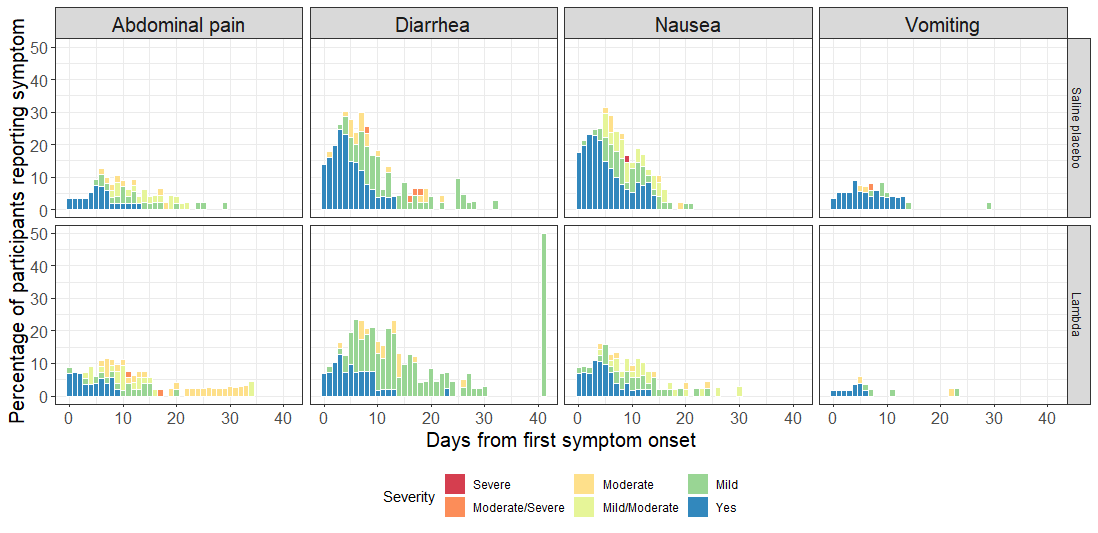

D. Other

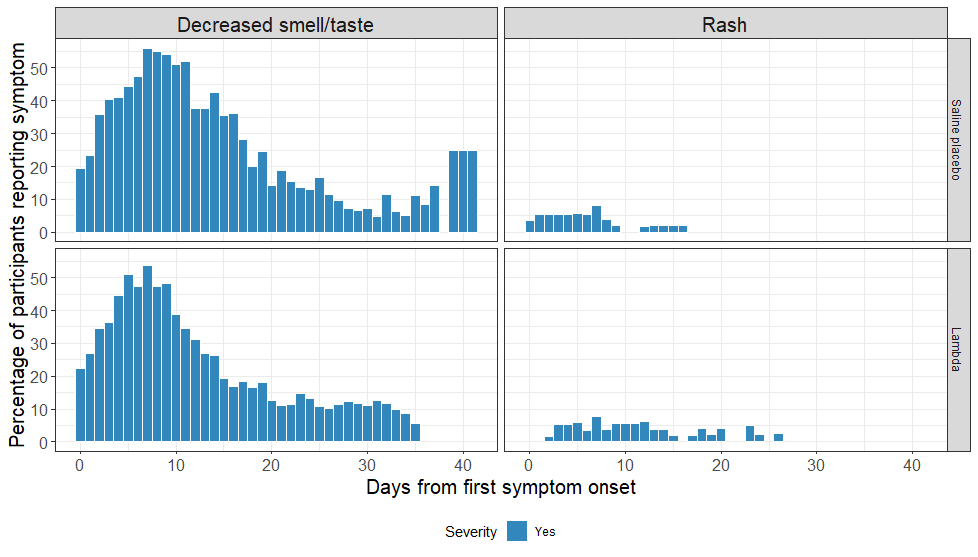

Figure S4: Distribution of the percentage of participants reporting each symptom by day from symptom onset and treatment arm, broken down by organ system. Color represents the severity of the symptom reported.

Figure S5. Pairwise correlation plots of symptoms by days from symptom onset

A. Day 0

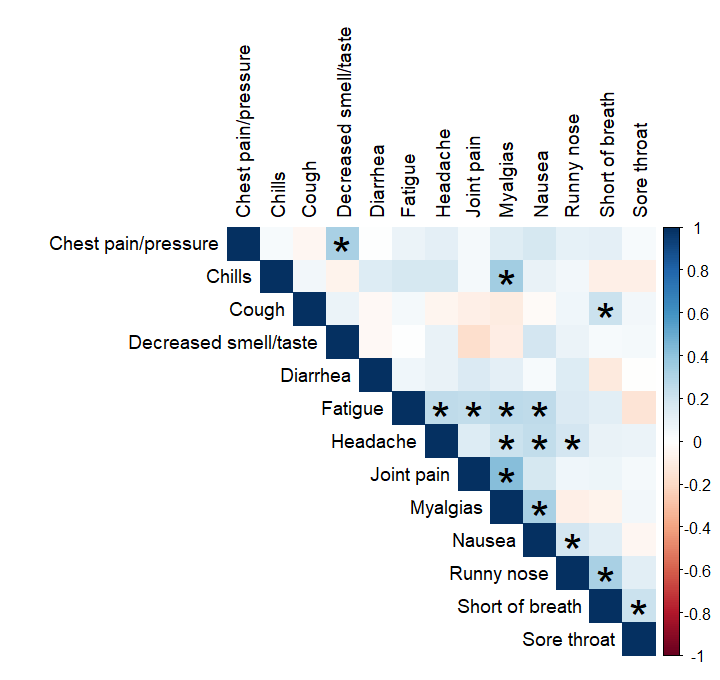

B. Day 4

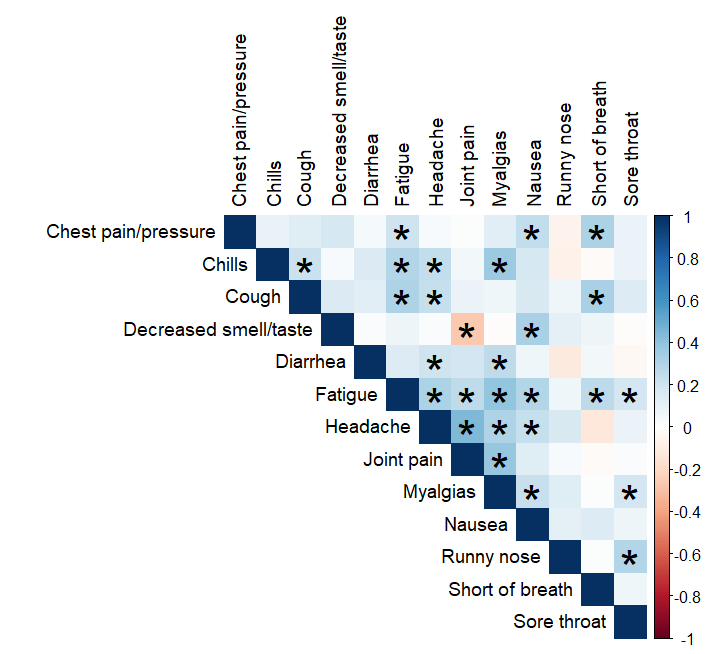

C. Day 7

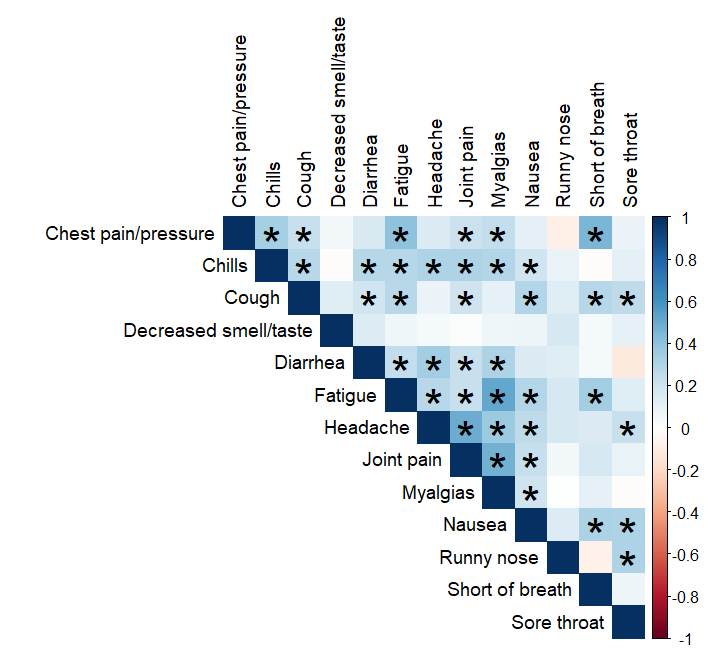

D. Day 10

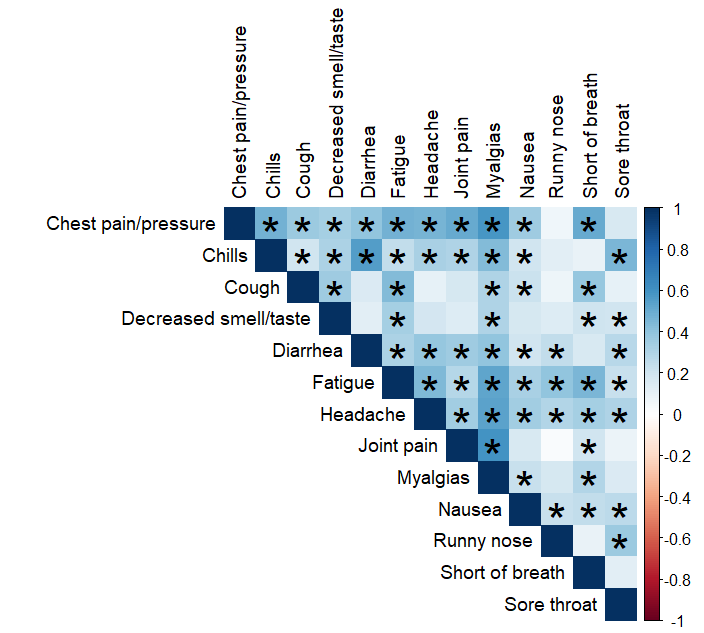

E. Day 14

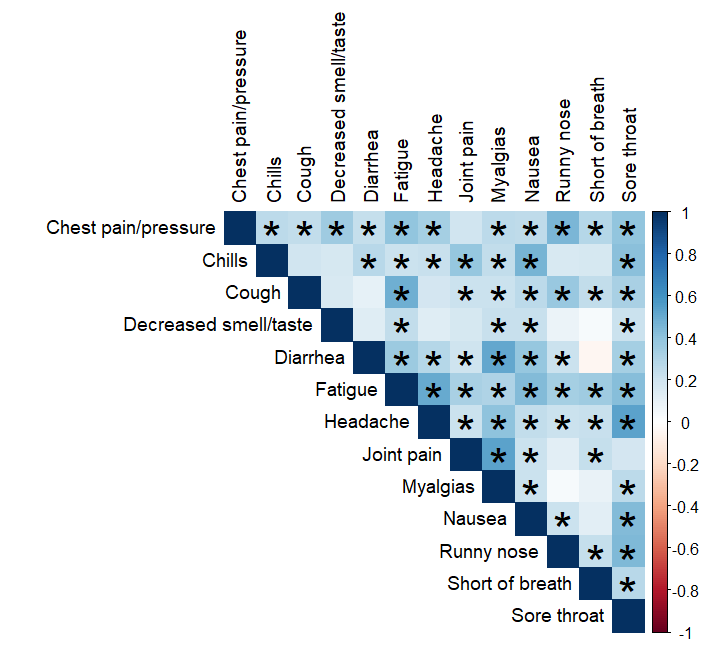

F. Day 21

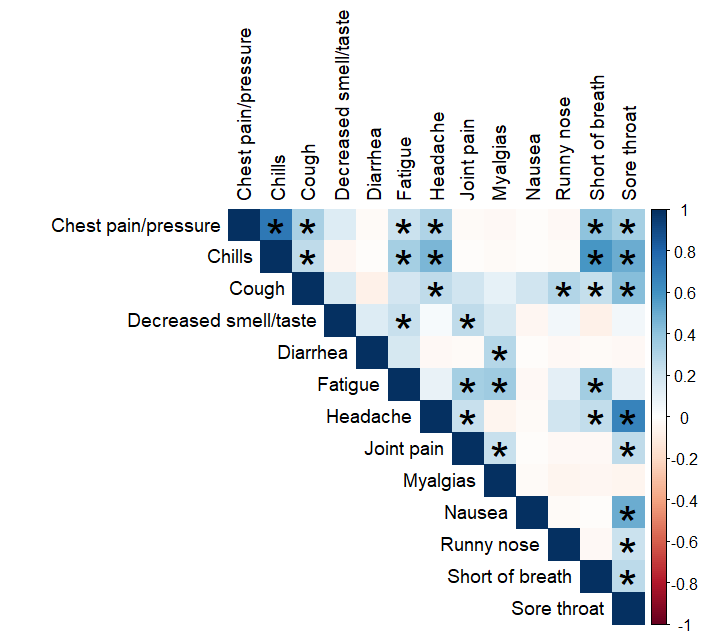

Figure S5: Correlation plots of symptoms at each day from symptom onset used in the exploratory factor analysis. Color indicates direction of correlation (red = negative, blue = positive) while shading indicates magnitude of association (light = weak, dark = strong). Asterisks represent statistically significantly correlations based on Spearman’s rank correlation co
